# Bacteriological Contamination of Shared Bicycle Handlebars and Hand Hygiene Practices of Users at Makerere University, Kampala, Uganda

**DOI:** 10.64898/2026.11.10.26367775

**Authors:** Catherine Daniella Tendo, James Muleme, Abdullah Ali Halage, Raymond Nicholas Mwaka, Caroline Nabagala, Maxentia Walusimbi, Charles Ssemugabo

## Abstract

**Background:** Bacteriological surface contamination is a key pathway for pathogen transmission, with potential implications for microbial exposure. Shared bicycles at Makerere University provide sustainable transport, but frequent handling of handlebars may facilitate colonization and spread of bacteria such as *E. coli.* This study aimed to identify prevalent bacterial species on shared bicycle handlebars, their microbial load and antimicrobial profiles, and users’ hand hygiene practices influencing contamination at Makerere University in Kampala, Uganda.

**Methods:** We randomly sampled 100 shared bicycles and interviewed their respective users. Handlebar swabs were cultured and analyzed by culture-based enumeration. Antimicrobial susceptibility testing was performed using disk diffusion across six antibiotics. Hand hygiene data were collected using structured questionnaire guides. Descriptive statistics summarized bacterial counts, F-tests assessed statistical differences within and across bacterial species, and modified poisson regression examined associations between hygiene practices and contamination.

**Results:** Contamination was detected on 56% (56/100) of the sampled handlebars. From the 56 contaminated samples, 13 contained more than one bacterial species and 75 isolates were recovered, with *S. aureus* at 68% (51/75) as the most prevalent isolate. Microbial loads differed significantly across species (F (2, 63) = 12.78, p < 0.0001). 35 of the 75 isolates yielded bacteriological growth and were subjected to AST. Most isolates were resistant to erythromycin at 71% (25/35), and *S. aureus* showed resistance to five of six antibiotics, indicating multidrug resistance. Users who did not clean their hands were (APR = 1.61, 95% CI: 1.11–1.98) 1.6 times more likely to contaminate handlebars.

**Conclusion:** Shared bicycle handlebars at Makerere University were contaminated with potentially pathogenic bacteria, indicating a potential hygiene concern. Not cleaning hands was associated with contamination of bicycle handlebars. The service provider urgently needs to promote proper hand hygiene by installing hand-washing stations with soap or sanitizers at docking points

## Introduction

Globally, universities are prioritizing the promotion of physical activity through the integration of sustainable mobility measures (1). For instance, Sapienza University in Rome implemented a Sustainable University Mobility Plan (SUMP), which integrated bicycle infrastructure like designated cycling lanes for bicycle riders as well as walking paths for pedestrian mobility, and charging stations for electric fleet transition to reduce reliance on fuel vehicles that cause environmental pollution (2). The introduction of shared bicycles in Makerere University, Uganda, has provided an environmentally friendly and affordable transport option (3). However, the frequent and communal use of these shared bicycles raises concern about surface hygiene, as handlebars serve as high-contact points that may harbor pathogenic microorganisms (4).

The surfaces of shared modes of transport are hotspots for various biological hazards (5) and serve as reservoirs for pathogens (6) facilitating their transmission among the different users by contact (7). Bacterial contamination has been documented on public means of transport like buses (7), trains (8), bicycles (9). Individuals who use communal services of movement unknowingly expose themselves to pathogenic bacteria like *E. coli*, *Vibrio cholerae*, and multidrug-resistant *S. aureus* (4). This poses a potential threat to public health, as there is an increased risk of the spread of infections caused by these microbial species (10) such as typhoid, gastroenteritis, and wound sepsis (11) that can lead to mortality if not promptly managed (12).

Bacteriological contamination in Uganda on public surfaces has been documented on paper currency, where *S. aureus* was found to be dominant (13). These findings highlight the risk of cross-contamination during cash transactions in shared mobility systems. Makerere University in Kampala, Uganda, adopted the bicycle sharing program in 2023 to facilitate mobility of the staff, students, and visitors around the 350-acre campus. Makerere University, with an estimated population of 40,000 individuals, operates approximately 120 shared bicycles, yielding a user-to-bicycle ratio of 1:300, underscoring handlebars as frequently touched surfaces. The initiative supports sustainable mobility but also raises public health concerns, as frequent handlebar handling combined with cash payments increases the potential for microbial exposure. Therefore, this study aimed to understand the extent of bacteriological contamination on the handlebars and hand hygiene practices of the shared bicycle users at Makerere University, Kampala, Uganda.

## Methods

### Study settings and design

This cross-sectional study was conducted at Makerere University, a public institution in Kampala, Uganda, between June and August 2025 using quantitative data collection methods. The sustainable mobility program at Makerere University has twelve docking stations, but only eight were actively functional, with bicycles used by students, staff, and visitors at the time of data collection.

Makerere University was selected because it is the only institution with a bicycle-sharing program, offering a unique context for study. Shared bicycles were chosen over other public means of transport like taxis and motorcycles since the user holds the handlebars, which involves repeated hand contact by multiple users, making them particularly susceptible to microbial contamination.

### Study participants

This study was carried out among bicycle users and shared bicycles at Makerere University. Eligible participants were students, staff, and visitors who were actively using the shared bicycles during the study period and consented to provide hand hygiene information. Only shared bicycles in routine circulation and accessible at docking stations were included for sampling.

### Sample size and Sampling

The study used 100 shared-bicycle users and their corresponding ridden bicycles. At docking stations, multiple users often retrieved bicycles concurrently. Swab collection from the bicycle handlebars and interviews were conducted on one participant at a time whose trip had ended. The users were present, and recruitment was contingent on participant consent, while individuals who declined were not included in the study. The Kish and Leslie (1965) (14) formula was used. Considering a Z-score (95% confidence interval) of 1.96, the prevalence of contamination of bicycle handlebars with bacteria of 50%, and a degree of error of 5%, 384 participant/bicycles will be studied. We adjusted for a finite population of 120 shared bicycles using Cochran’s (1977) formula, generating a total of 92. The minimum required sample size after finite population correction was 92. However, laboratory analysis was conducted on a rolling basis, and some collected samples did not yield bacterial growth. To compensate for these losses and ensure sufficient analyzable data, we modestly increased recruitment to 100 participants, increasing the number of observations available for analysis of the study.

### Data collection

Surface swab samples were collected from bicycle handlebars and transported to the Microbiology Diagnostic Laboratory (Biosafety level-2) in Makerere University for analysis. Subsequently, a structured questionnaire was used to collect data on the bicycle users’ hand hygiene practices. Each bicycle and the respective user were both assigned identical study identification numbers to enable merging of questionnaire responses and the bacteriological profiles found on the bicycle.

Handlebar swab samples were collected with sterile, wet cotton swabs of 15mm (15). The ends of the swabs were aseptically broken to fit in a Falcon tube (Conda Lab, Spain) of 15ml capacity with a screw cap and conical bottom containing 5ml of Stuart’s transport medium (Hi Media Laboratories Limited, India). They were placed in a cooler box and then transported to the laboratory for analysis.

### Sample processing

In the laboratory, the swabs were removed from the Falcon tube using sterile forceps and placed in a standard test tube of 20mL containing 9mL of sterile peptone water (Conda Lab Madrid, Spain). The primary bacteriological suspension was labelled 10^-1^ and agitated for about 30 seconds using a pipette tip to extract bacteria from the swab. This was followed by a 10-fold serial dilution to get 10^-2^ and 10^-3^ solutions. These were separately inoculated and surface-spread onto sterile Mannitol Salt Agar plates (MSA) and MacConkey Agar plates (MAC), both manufactured by Hardy Diagnostics in Santa Maria, U.S.A. The plates were placed in an incubator (Memmert, Germany) at 37 °C for MSA and 45 °C for MAC for about 20 hours. Microbial load was then calculated as CFU=N/df x V, where N=number of colonies, df=dilution factor (10^-3^), V=volume plated (10^-1^), and expressed as log CFU/mL× 10². Subculturing was done to obtain pure colonies of the isolates (Muhumuza et al., 2018).

The presumptive bacteriological pure colonies identified by their morphology and color that is pink for coliforms and yellow for *S. aureus* were then subjected to standard biochemical tests for confirmation (17,18). *E. coli* suspected isolates were examined using Sulfide Indole Motility (S.I.M), Triple Sugar Iron (T.S.I), Methyl Red-Voges-Proskauer, urease, and citrate (19). *S. aureus* suspected isolates were confirmed by catalase testing and Gram staining (20). Antimicrobial susceptibility testing (AST) was performed using the Kirby-Bauer disk diffusion method, and interpretation of the results as susceptible, intermediate, and resistant (S/I/R) was carried out based on the standard reference criteria provided by the 2024 CLSI guidelines (21). Subculturing was done to obtain pure cultures of the isolates, from which discrete colonies were emulsified into a test tube with 1 mL of normal saline. The bacteriological suspension was compared to a standard of 0.5 McFarland to compare the optical density. If the turbidity matched, then a sterile cotton swab was immersed in the suspension and used to inoculate Mueller-Hinton Agar (MHA) plates by surface spreading.

Antibiotic disks (Hardy Diagnostics, U.S.A) were applied to the MHA plates using sterile forceps. The choice of antibiotics was obtained from Clinical and Laboratory Standards Institute (CLSI) guidelines (22), these were Amoxicillin-clavulanic (AMC32 µg), Gentamicin (CN120 µg), Ciprofloxacin (CIP5 µg), Cefoxitin (FOX30 µg), Imipenem (IPM10 µg), Sulfamethoxazole-Trimethoprim (SXT25 µg), Erythromycin (E15 µg), and Tetracycline (Te30 µg). The plates were incubated at 37°C for 24 hours, after which the diameter of the clear zone of inhibition was measured and documented.

Users’ hand hygiene data were obtained using structured questionnaires administered to the participants electronically with Kobo Collect. The questionnaire was developed from literature on hand hygiene and microbial contamination (23,24). The tool was structured into four sections on demographics, bicycle usage patterns, hygiene practices, and additional information on contamination perceptions. The questionnaire guide was refined through expert input by supervisors and pre-testing. The pre-testing of the tool was carried out amongst 9 classmates that are shared bicycle users at Makerere University to assess the relevance, clarity, and suitability of the questions. Feedback obtained during the pre-test was used to refine the questionnaire before the main data collection. The classmates who participated in the pre-test were excluded from the final study sample. Trained research assistants administered the interviews face-to-face at docking stations, approaching users immediately after parking their bicycles to capture real-time responses.

### Data analysis

Data were analyzed using STATA software version 18. Descriptive statistics was used to summarize the socio-demographic characteristics of participants and to present the distribution of hand hygiene practices and levels of bacteriological contamination observed on bicycle handlebars. One-way ANOVA with F-tests was used to compare mean microbial loads across bacterial species, to determine whether contamination intensity differed significantly between isolates. In contrast, chi-square tests were applied to assess associations between categorical predictors e.g., hand hygiene practices, age group, frequency of bicycle use and contamination status. Modified poisson regression was used to estimate the prevalence ratios and their 95% confidence intervals to quantify associations between bacteriological contamination and hand hygiene practices while adjusting for potential confounders (e.g., age, sex, frequency of bicycle use, college affiliation, and access to hygiene facilities).

### Ethical consideration

Ethical approval was obtained from the Makerere University School of Public Health Higher Degrees Committee, and the study protocol number with MakSPH-REC was 599. Permission was granted by Makerere University administration and the shared bicycle service provider to conduct the study formally within institutional guidelines and with the cooperation of the service operators. Written informed consent was obtained from all study participants before bicycle handlebar sampling and questionnaire administration.

Confidentiality and anonymity were strictly maintained throughout the study. Anonymity was ensured by assigning unique codes to participants instead of recording personal identifiers such as names or contact details. All responses were stored in password-protected files on the principal investigator’s computer, accessible only to the research team.

Data were used solely for research purposes and reported in aggregate form, preventing identification of individual participants. Biosafety protocols were rigorously observed during sample collection and laboratory analysis, including the use of personal protective equipment, sterilization of materials, and adherence to standard operating procedures, thereby safeguarding researcher safety and maintaining sample integrity.

## Results

### Socio-demographic characteristics of the study participants

A total of 100 respondents participated in this study; of these, 55% were male, and 82% were between 18 and 25 years of age. Frequency of bicycle use varied, with 47% reporting daily use as shown in the **Table 1**.

**Table 1:**
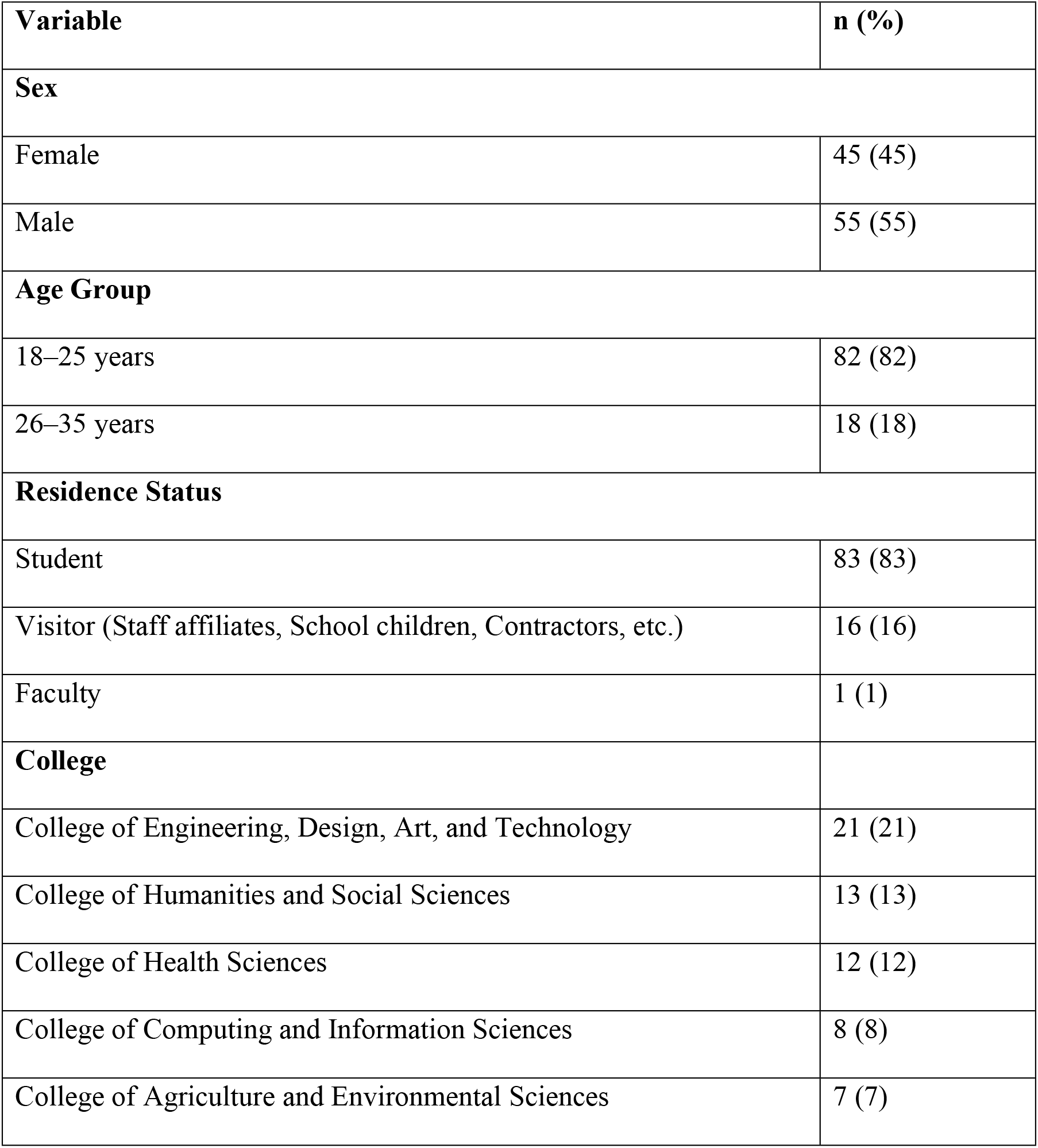
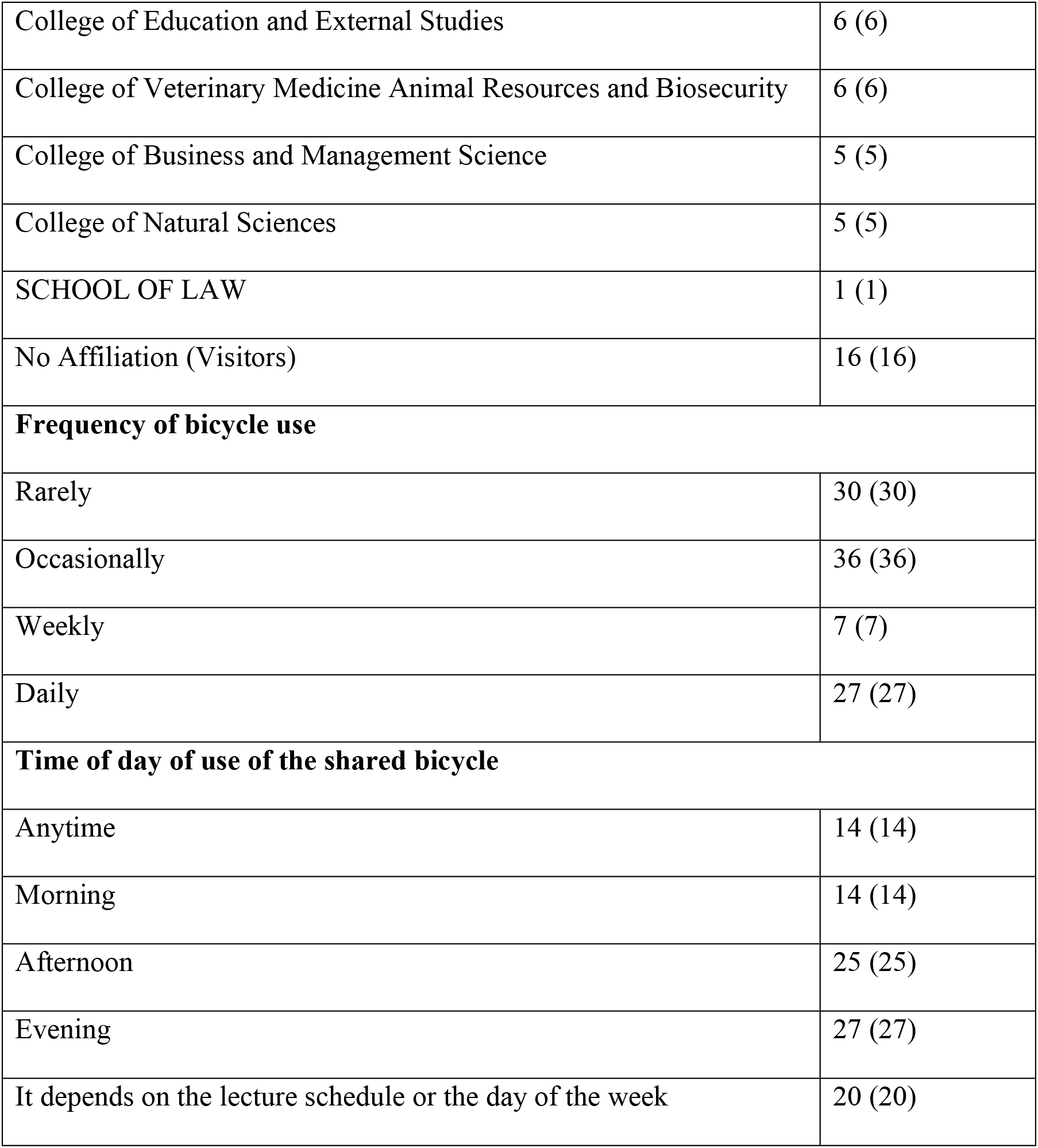
Socio-demographic characteristics of study participants.

| **Variable** | **n (%)** |
|---|---|
| **Sex** |  |
| Female | 45 (45) |
| Male | 55 (55) |
| **Age Group** |  |
| 18–25 years | 82 (82) |
| 26–35 years | 18 (18) |
| **Residence Status** |  |
| Student | 83 (83) |
| Visitor (Staff affiliates, School children, Contractors, etc.) | 16 (16) |
| Faculty | 1 (1) |
| **College** |  |
| College of Engineering, Design, Art, and Technology | 21 (21) |
| College of Humanities and Social Sciences | 13 (13) |
| College of Health Sciences | 12 (12) |
| College of Computing and Information Sciences | 8 (8) |
| College of Agriculture and Environmental Sciences | 7 (7) |
| College of Education and External Studies | 6 (6) |
| College of Veterinary Medicine Animal Resources and Biosecurity | 6 (6) |
| College of Business and Management Science | 5 (5) |
| College of Natural Sciences | 5 (5) |
| SCHOOL OF LAW | 1 (1) |
| No Affiliation (Visitors) | 16 (16) |
| **Frequency of bicycle use** |  |
| Rarely | 30 (30) |
| Occasionally | 36 (36) |
| Weekly | 7 (7) |
| Daily | 27 (27) |
| **Time of day of use of the shared bicycle** |  |
| Anytime | 14 (14) |
| Morning | 14 (14) |
| Afternoon | 25 (25) |
| Evening | 27 (27) |
| It depends on the lecture schedule or the day of the week | 20 (20) |

### Bacteriological identification, bacterial load and antimicrobial susceptibility profiles

Of the 100 samples analyzed, 56 were positive for bacteriological contamination, accounting for 56% of the overall bacteriological contamination of shared bicycle handlebars. As shown in **Table 2** from the 56 contaminated samples, a total of 75 bacterial isolates were recovered, as some samples yielded more than one bacterial species. Therefore, the prevalence of individual bacterial species was calculated using the total number of isolates recovered. *Staphylococcus aureus* was the predominant isolate, accounting for 68% of contaminated samples. Thermotolerant bacteria were detected in 17% of contaminated samples, while total coliforms were present in 15%, which may indicate fecal contamination.

**Table 2:** Identified bacteriological species and their prevalence.

| **Variable** | **Frequency (n)** | **Percentage (%)** |
|---|---|---|
| **Handlebar samples examined** | 100 | 100.0 |
| Contaminated | 56 | 56.0 |
| Not-contaminated | 44 | 44.0 |
| **Number of bacterial species per contaminated sample (n=56)** |  |  |
| One bacterial species | 43 | 76.8 |
| Two bacterial species | 9 | 16.1 |
| Three bacterial species | 4 | 7.1 |
| **Bacterial isolates recovered (n=75)** |  |  |
| *Staphylococcus aureus* | 51 | 68.0 |
| Thermotolerant bacteria | 13 | 17.3 |
| Total coliforms | 11 | 14.7 |

#### Differences in bacterial load among bacterial species

The F-test (F(2,63)=12.78, p<0.0001) demonstrated that mean contamination levels varied significantly across the groups. Post-hoc comparisons revealed that *S. aureus* had significantly higher contamination levels compared to total coliforms (mean difference=1.321 log CFU/mL, p<0.001) and thermotolerant coliforms (mean difference=0.462 log CFU/mL, p=0.121, not significant). The comparison between total coliforms and thermotolerant coliforms also showed a significant difference (mean difference=0.859 log CFU/mL, p=0.009). These results confirm that *S. aureus* was not only the most frequently isolated organism but also exhibited significantly higher contamination levels as presented in **Table 3**, underscoring its public health relevance.

**Table 3:**
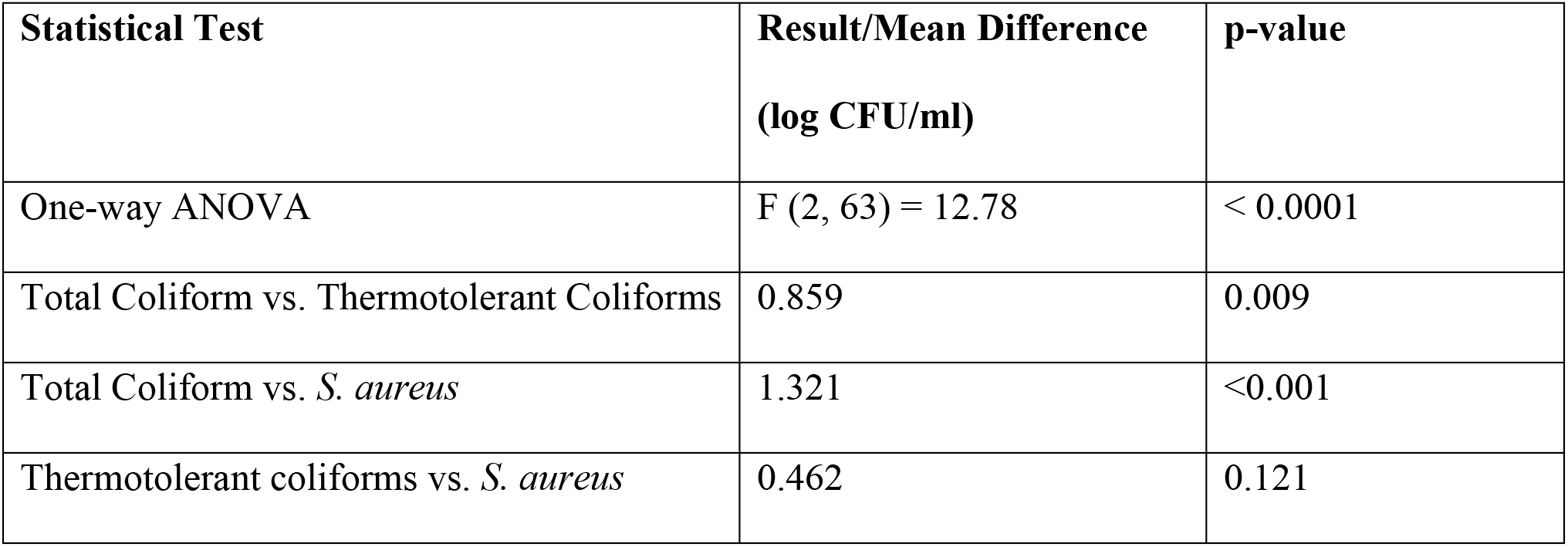
One-way ANOVA and pairwise comparison of bacteriological contamination levels.

| **Statistical Test** | **Result/Mean Difference (log CFU/ml)** | **p-value** |
|---|---|---|
| One-way ANOVA | F (2, 63) = 12.78 | < 0.0001 |
| Total Coliform vs. Thermotolerant Coliforms | 0.859 | 0.009 |
| Total Coliform vs. *S. aureus* | 1.321 | <0.001 |
| Thermotolerant coliforms vs. *S. aureus* | 0.462 | 0.121 |

#### Antimicrobial susceptibility tests to demonstrate species resistance profiles

Out of the 75 isolates from the 56 contaminated shared bicycle handlebar samples, 35 isolates yielded viable bacteriological growth and were subjected to confirmatory biochemical tests and thereafter subjected to antimicrobial susceptibility testing (AST). Of these, 25 (71.4%) were *S. aureus*, 9 (25.7%) were coliform bacteria, and 1 (2.9%) thermotolerant coliform was *E. coli*. AST revealed a high prevalence of resistance among the bacteriological isolates, particularly to Erythromycin (E15 µg) at 71%, with all total coliform isolates resistant at 100% (9/9). The *E. coli* (thermotolerant coliform) isolate was only resistant to Tetracycline (Te30 µg). Of the six antibiotics tested on *S. aureus,* it was resistant to five, underscoring the predominance of *S. aureus* as a resistant pathogen. The bacterial isolates did not show resistance to Amoxicillin-Clavulanate (AMC32 µg), Ciprofloxacin (CIP5 µg), and Imipenem (IPM10 µg), highlighting their continued therapeutic relevance as presented in **Figure 1**. The *S. aureus* isolates exhibited resistance to gentamicin (CN120 µg), trimethoprim-sulfamethoxazole (SXT25 µg), cefoxitin (FOX30 µg), erythromycin (E15 µg), and tetracycline (Te30 µg), representing resistance across five antimicrobial classes (aminoglycosides, folate pathway inhibitors, β-lactams, macrolides, and tetracyclines respectively), and was therefore classified the isolates as Multi Drug-Resistant (MDR) *S. aureus* further more resistant to cefoxitin (FOX30 µg), indicating methicillin resistance and therefore was classified as MRSA (25).

**Figure 1:**
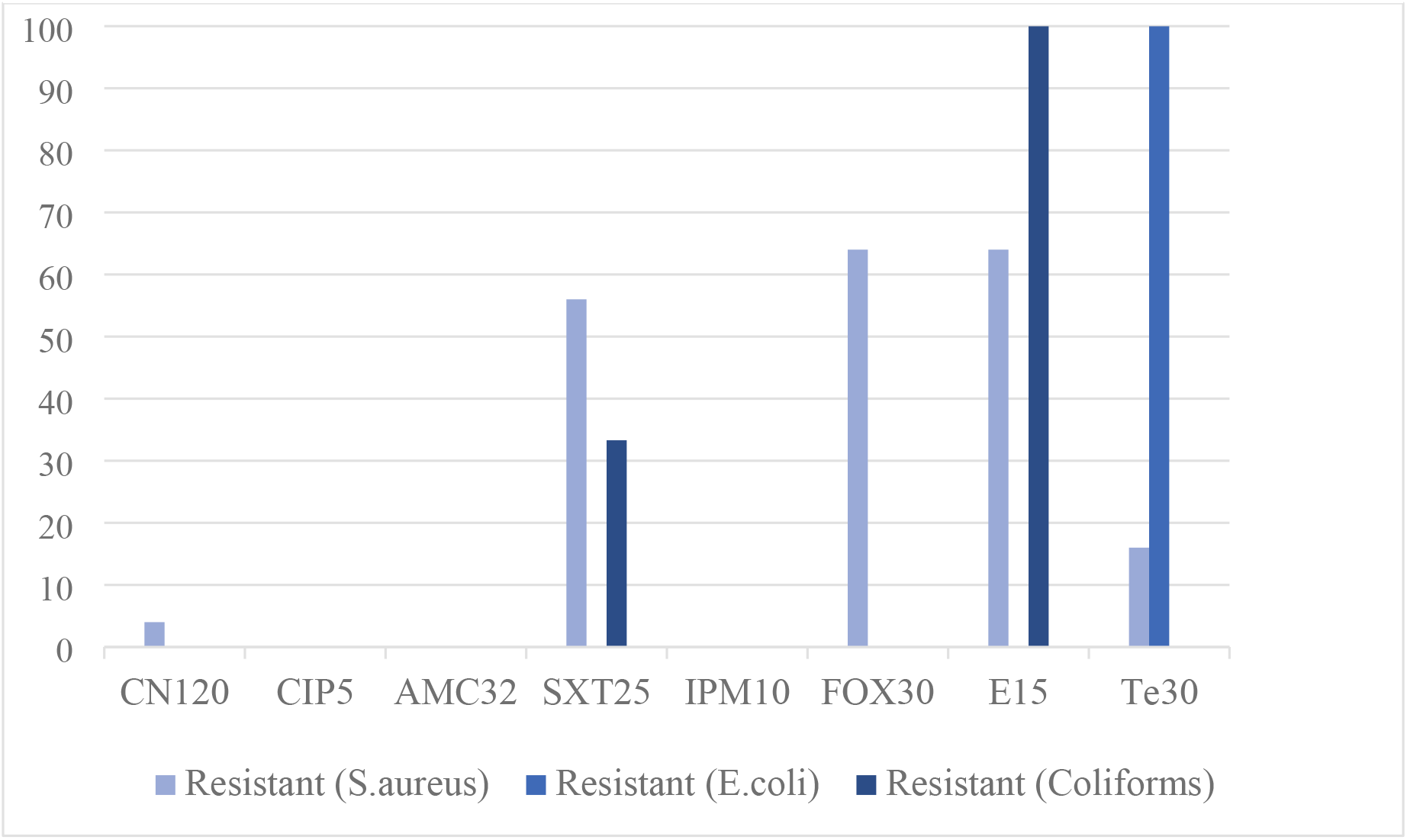
An antibiogram showing susceptibility of the isolates to antibiotics.

### Hand hygiene practices of the bicycle users

Only 25% (25/100) reported that they cleaned their hands before and after use as shown in **Table 4**. Among those who did not practice proper hand hygiene, most were not aware of proper hand hygiene practices at 43% (32/75), lacked access to hand hygiene facilities at 27% (20/75), and believed that their hands were not dirty at 16% (12/75). Of the 25 participants who practiced hand hygiene, most relied on sanitizers 60% (15/25), and used soap and water at 28% (7/25) or both methods at 12% (3/25).

**Table 4:** Hand hygiene practices of the users of the shared bicycles.

| Variable on hand hygiene | Category | Frequency (n) | Percentage (%) |
|---|---|---|---|
|  | Yes | 25 | 25.0 |
| Practicing hand hygiene before and after using the shared bicycles | No | 75 | 75.0 |
| Reasons for not practicing proper hand hygiene<br>(n=75) | Lack of awareness/consideration | 32 | 42.7 |
|  | No access to hand hygiene facilities | 20 | 26.7 |
|  | Perceive it as unnecessary | 5 | 6.7 |
|  | Too busy/in a hurry | 4 | 5.3 |
|  | Forgetfulness | 2 | 2.7 |
|  | Believe hands are not dirty | 12 | 16.0 |
| Hand hygiene protocol used (n=25) | Sanitizing | 15 | 60.0 |
|  | Handwashing with soap and water | 7 | 28.0 |
|  | Both methods | 3 | 12.0 |

#### Factors associated with bacteriological contamination of shared bicycle handlebars

Participants who did not clean their hands before and after use of shared bicycles had a 61% higher prevalence of contamination compared to those who did, even after adjustment (Adjusted PR=1.61, 95% CI: 1.11–1.98) as presented in **Table 5** below where (*) is statistically significant at p < 0.05.

**Table 5:** Factors associated with bacteriological contamination of shared bicycle handlebars.

| **Factor** | **Not Contaminated [n (%)]** | **Contaminated [n (%)]** | **Unadjusted PR [95% CI]** | **Adjusted PR [95% CI]** |
|---|---|---|---|---|
| **Sex** |  |  |  |  |
| Female | 27 (48.2%) | 29 (51.8%) | 1.00 (ref) | 1.00 (ref) |
| Male | 17 (38.6%) | 27 (61.4%) | 1.19 [0.87–1.62] | 1.25 [0.90–1.74] |
| **Age group** |  |  |  |  |
| 18–25 years | 37 (45.1%) | 45 (54.9%) | 1.00 (ref) | 1.00 (ref) |
| 26–35 years | 7 (38.9%) | 11 (61.1%) | 1.16 [0.78–1.47] | 1.13 [0.68–1.48] |
| **Cleaning of hands before and after use of a shared bicycle use** |  |  |  |  |
| Yes | 28 (57.1%) | 21 (42.9%) | 1.00 (ref) | 1.00 (ref) |
| No | 16 (29.6%) | 38 (70.4%) | **1.64 [1.21–1.96] \*** | **1.61 [1.11–1.98] \*** |
| **Frequency of bicycle use** |  |  |  |  |
| Occasionally (≤2 times/week) | 23 (56.1%) | 18 (43.9%) | 1.00 (ref) | 1.00 (ref) |
| Frequently (≥2 times/week) | 21 (33.9%) | 41 (66.1%) | 1.51 [1.07–1.85] | 1.44 [0.95–1.84] |

## Discussion

This study aimed at identifying the extent and nature of bacteriological contamination on shared bicycle handlebars. The handlebars were found to be contaminated with *S. aureus*, which was the most prevalent isolate, thermotolerant coliforms like *E. coli*, and total coliforms. Antimicrobial resistance testing revealed resistance to Erythromycin (E15 µg) amongst most isolates. Importantly, participants who did not wash their hands with soap demonstrated a higher prevalence of contamination, underscoring the critical role of soap use in reducing bacterial transfer.

The findings reveal relatively high bacteriological contamination of shared bicycle handlebars, with most of the bicycle handlebars sampled testing positive for at least one bacterial organism. Bacteria could be introduced onto surfaces through various means, including hand contact and environmental sources like wind (26). The level of contamination indicates potential for microbial exposure in community mobility systems, pointing to hygiene gaps in shared transport infrastructure. These results are consistent with findings from Ghana, where (8,27) reported bacterial contamination on shared public transport seat materials, suggesting that such contamination is a recurring challenge across different communal transport settings. This reinforces the notion that shared mobility surfaces are critical points of exposure and require targeted interventions. The service providers should implement routine cleaning and disinfection protocols, alongside public awareness campaigns on hand hygiene, to reduce contamination risks and safeguard public health.

Among the bacterial species isolated, *S. aureus* was the most predominant, followed by total coliforms and *E. coli*. The predominance of *S. aureus* on the bicycle handlebars may reflect its common colonization of human skin and nasal passages and its potential transfer through hand contact. The detection of *E. coli* and total coliforms, albeit at lower prevalence, is of particular public health concern as they are indicators of faecal contamination (7). Shared bicycles are high-touch surfaces frequently used by multiple individuals in succession, which creates an ideal environment for the persistence and spread of bacteriological organisms. Sun et al., (2020) (28) reported *Bacillus* species as the dominant contaminants on bicycle seats and handlebars, suggesting that differences in environmental conditions, user populations, and hygiene practices may influence the dominant bacterial species across settings. The findings highlight the importance of routine cleaning/disinfection of handlebars and the promotion of hand hygiene practices among users to reduce opportunities for the risk of bacterial transmission The antimicrobial susceptibility testing revealed widespread resistance patterns, with *S. aureus* resistant to most of the antibiotics tested, coliforms showing consistent resistance to erythromycin, and *E. coli* exhibiting selective resistance to tetracycline while retaining sensitivity to amoxicillin-clavulanate, ciprofloxacin, and imipenem. Prior investigations have also reported the prevalence of pathogenic microorganisms on surfaces, such as methicillin-resistant *S. aureus* (13,26). This finding signifies the growing challenge of antimicrobial resistance in community environments, where resistant strains can persist on high-touch surfaces and increase the risk of transmission, which calls for stricter regulation of antibiotic use to curb misuse and integration of sanitation protocols in shared mobility systems to minimize exposure to resistant strains. Ugandan studies have also documented extended-spectrum beta-lactamase-producing *E. coli* (ESBL) and antimicrobial-resistant *E. coli* across human, animal, and environmental compartments, supporting the importance of community and environmental reservoirs in AMR surveillance (30,31).

The study found considerable variation in hand hygiene practices among shared bicycle users, with only a minority reporting consistent hand cleaning after trips. Among those who practiced hygiene, the majority relied on sanitizers, while a smaller proportion used soap and water, typically accessed in lecture rooms or places of residence. The reported use of sanitizers may reflect convenience; however, this study did not compare the effectiveness of sanitizing with soap-and-water handwashing. These findings are consistent with broader public health literature, which often documents a disconnect between knowledge of hygiene practices and their implementation in daily routines (29,30). These findings underscore the importance of integrating hygiene promotion into shared mobility systems, including embedding hand hygiene awareness campaigns within university curricula and health clubs to encourage consistent hand hygiene practices among students and staff.

The study demonstrated a significant association between hand hygiene practices and the level of bacterial contamination on shared bicycle handlebars, with inadequate handwashing linked to a higher prevalence of contamination (34). This finding signifies that behavioral factors play a critical role in shaping microbial risks in shared mobility systems, reinforcing the importance of user practices alongside environmental conditions. (35) reported a positive correlation between frequency of public bicycle use and contamination burden, likely due to repeated inoculation events from multiple users. The findings suggest that shared bicycles, while beneficial for sustainable urban transport, may act as high-touch surfaces on which bacteria can be detected and potentially transferred, particularly if hygiene practices are suboptimal. Behavioral factors such as neglecting hand hygiene or failing to use soap play a critical role and represent modifiable risk factors. This underscores the need for behavioral change interventions like digital nudges, such as app-based reminders that prompt users to clean their hands at the end of the trip after docking bicycles. Targeted health promotion campaigns emphasizing regular hand hygiene (36,37) and disinfection of shared bicycle handlebars (38) could lower microbial load and interrupt transmission pathways.

The study had strengths and limitations. This study provides valuable insights into the bacteriological contamination of shared bicycle handlebars at Makerere University, highlighting the presence of pathogenic bacteria, including multidrug-resistant *Staphylococcus aureus*. These findings contribute novel evidence from a sub-Saharan university setting, where research on microbial risks in shared mobility systems remains scarce.

However, the relatively small sample size of 100 constrained the testing of associations between contamination and hand hygiene behavior, limiting the statistical significance of observed relationships. In addition, the study population was drawn from a university community that is relatively closed and more educated compared to the broader users of public transport in Uganda, restricting the generalizability of findings to diverse settings such as taxis and buses. Another limitation of the study was that only bicycle handlebar samples were collected, while no hand-rinse or hand-swab samples were obtained from participants because the bacteriological contaminants recovered from handlebars might have originated from previous users, not necessarily the one interviewed, or environmental sources. Despite these limitations, the study offers important data to inform hygiene interventions and antimicrobial resistance surveillance in communal transport environments.

## Conclusion

This study demonstrated that shared bicycle handlebars at Makerere University are contaminated with multiple bacterial species, most notably *Staphylococcus aureus*, total coliforms, and *Escherichia coli*, showing that these communal surfaces can carry potentially pathogenic organisms. Antimicrobial susceptibility testing revealed widespread resistance, including multidrug-resistant *S. aureus*, highlighting the relevance of AMR surveillance beyond clinical settings.

Statistical analysis identified an association between contamination levels and hand hygiene practices, with inadequate hand cleaning before and after use of the shared bicycles associated with a higher prevalence of contamination. These findings signify that both environmental conditions and user behaviors jointly shape microbial persistence and transmission in shared mobility systems. Consistent with earlier literature on public transport and other high-contact communal surfaces, our results highlight the urgent need for integrated interventions.

The university administration and the bicycle service provider should prioritize routine cleaning/disinfection of handlebars and other high-touch bicycle surfaces, expand access to hand-washing facilities near docking stations, and implement awareness campaigns that emphasize proper hygiene practices. In addition, surveillance of antimicrobial resistance in community settings should be strengthened to inform antibiotic stewardship and safeguard public health.

## Data Availability

The data set contains potential identifying information and thus to protect the participants privacy the data can not be made publicly available. Data are however made available upon request from the research and ethics committee by

## Acknowledgements

The author sincerely appreciates the guidance, mentorship, and constructive feedback provided by the academic staff and colleagues at Makerere University. Appreciation is also extended to the study participants and the bicycle-sharing company staff for their time and willingness to take part in the study.

